# Transcranial Focused Ultrasound Targeting the Default Mode Network for the Treatment of Depression

**DOI:** 10.1101/2024.05.16.24307494

**Authors:** Jessica N. Schachtner, Jacob F. Dahill-Fuchel, Katja E. Allen, Christopher R. Bawiec, Peter J. Hollender, Sarah B. Ornellas, Soren D. Konecky, Achal S. Achrol, John J.B. Allen

## Abstract

**Importance:** Up to 50% of individuals fail to respond to current depression treatments. Repetitive negative thought and default mode network hyperconnectivity are central in depression and can be targeted using novel neuromodulation techniques.

**Objective:** This study assessed whether non-invasive transcranial focused ultrasound to the default mode network can decrease depression symptoms and repetitive negative thought, and improve quality of life.

**Design:** This open-label case series began in August 2023, with a six-month follow-up period (current).

**Setting:** A community-based study at the University of Arizona.

**Participants:** Twenty individuals aged 18 – 45 were enrolled from among 247 screened. Exclusion criteria included history of psychosis/mania, acute suicidality, MRI contraindications, pregnancy, and medical and neurological factors that may complicate diagnosis or brain function.

**Intervention:** Up to three weeks of transcranial ultrasound (11 sessions) targeting the anterior medial prefrontal cortex; ten minutes per session.

**Main Outcomes and Measures:** Depression severity (Beck Depression Inventory – II and the Hamilton Depression Rating Scale), repetitive negative thought (Perseverative Thinking Questionnaire), and quality of life (World Health Organization Quality of Life survey) were outcomes.

**Results:** This sample was young (mean 30.4 years ± 10.0), predominantly female (75%), with moderate to severe depression and high comorbidity. Fifty percent of participants endorsed current psychiatric medication use. Ten percent of subjects dropped out of the study. Significant decreases in depression occurred on self-report, 11.3 (p < 0.001, CI = −14.68, −8.15) and interview ratings, 4.3 (p < 0.001, CI = −6.21, −2.43). Repetitive negative thought decreased by 8.53 (p <0.001, CI = −11.01, −5.79). Physical and psychological well-being improved by 7.6 (p < 0.001, CI = 3.62, 11.63) and 11.9 points (p < 0.001, CI = 7.51, 16.21), respectively. Environment satisfaction increased by 5.0 (p = 0.001, CI = 2.24, 7.56).

**Conclusions and Relevance:** Transcranial ultrasound holds promise as a treatment for depression.

**Trial Registration:** Altering Default Mode Network Activity with Transcranial Focused Ultrasound to Reduce Depressive Symptoms (DMNtFUS). Registration number: 019782-00001 Clinical trials ID: NCT06320028 URL: https://clinicaltrials.gov/study/NCT06320028?intr=Ultrasound&cond=depression&locStr=Arizona&country=United%20States&state=Arizona&rank=1

## Introduction

Depression is a leading cause of disability ^1^, affecting 21 million adults and significantly diminishing quality of life ^2^. Major Depressive Disorder (MDD) is typically recurrent ^3–5^, and impairment is compounded with subsequent episodes ^6^. Critically, current interventions are not effective for certain profiles of depression ^7,8^.

In conjunction with depressed mood and related symptoms, Repetitive Negative Thought (RNT) has been identified as a maintaining factor in depression^9^, as well as a predictor of depression improvements^8^. The brain’s Default Mode Network (DMN), which has greater connectivity during self-referential processing (e.g., mind-wandering ^10,11^) and, in particular, *negative* self-referential processing (e.g., RNT ^12^), is also shown to play an important role in depression.

Studies have identified that greater DMN connectivity (e.g., hyperconnectivity) has been associated with greater depression severity and RNT ^13,14^. Together, these findings highlight the mechanistic roles that RNT and DMN hyperconnectivity play in the development and maintenance of depression.

Because roughly 50% of depressed individuals are treatment-resistant to traditional treatments^7,15^, more effective interventions are needed, ideally those deriving from a better mechanistic understanding of depression. DMN connectivity has been altered (e.g., using transcranial magnetic stimulation (TMS), psychedelics, meditation) in various clinical populations ^16,17^, with the goal of improving treatment approaches. A novel neuromodulation technique, non-invasive Transcranial Focused Ultrasound Stimulation (tFUS), holds promise in the treatment of depression ^18,19^.

Unlike other noninvasive methods (TMS and transcranial electrical stimulation (TES) using direct (tDCS) or alternating (tACS) current), tFUS uses low-intensity ultrasound involving a focused nonthermal ultrasound beam, which safely passes through the skull ^20^ to exert electro-mechanical effects on target neurons, including the ability to induce excitatory and inhibitory effects depending on the sonication parameters used ^21,22^. tFUS also presents advantages beyond other non-invasive neuromodulation techniques (e.g., TMS) due to its ability to target deeper brain regions with greater precision ^22^, without side effects (e.g., skin irritation, local pain) that can accompany techniques like TMS ^23^.

Limited research supports tFUS as a treatment for depression. Resnik and colleagues examined transcranial ultrasound targeting the right inferior frontal gyrus, a component of the executive control network, on symptoms of depression; those engaging in a five-day treatment regime experienced a reduction in worry ^18^ compared to those receiving sham. Additionally, Sanguinetti and colleagues also found that tFUS reduced negatively-valanced emotions and altered DMN connectivity ^19^. These findings provide the foundation for further exploring the use of tFUS as a treatment for depression.

The present study aimed to assess whether tFUS delivered to the anterior medial prefrontal cortex (amPFC), a hub of the DMN^11^, can reduce depression symptoms and RNT, improve quality of life, and whether changes in depression severity are mirrored by changes in RNT.

## Methods

The Institutional Review Board of the University of Arizona approved the experimental protocol. All participants signed an informed consent document before participation.

### Participants

Individuals with a current major depressive episode, assessed using the Structured Clinical Interview for the DSM-5, were enrolled. They also experienced clinically significant RNT, characterized by a total score on the Perseverative Thinking Questionnaire (PTQ) ^24^ above the 75% percentile (≥37). Participants were ages 18 – 45, right-handed, English-speaking, and without any neurological symptoms or symptoms of mania/psychosis. Additional exclusion criteria included: history of head injury with loss of consciousness; uncorrected vision and/or hearing impairment that would interfere with study participation; current or history of brain or mental illness judged likely to interfere with testing, including drug and/or alcohol dependence; a diagnosed sleep disorder (e.g., Insomnia); current drug, alcohol or prescription drug intoxication; history of epilepsy; history of diagnosed migraines; metal implants in head or face, including permanent dental retainers; history of cardiac problems that could impact brain function (e.g., atrial fibrillation); and current active suicidal ideation necessitating immediate treatment.

### Overview of ultrasound treatment protocol

Eligible participants completed up to three weeks of ultrasound treatment. Before treatment, they completed an MRI session, a clinical interview, and self-report surveys. The first week of ultrasound involved five sessions within a seven-day period. Participants completed the same baseline assessments after completing week one, and if they did not meet early remission criteria (defined below), they continued ultrasound treatment for two more weeks, three sessions per week, each within a seven-day period. Participants completed the same series of assessments after week three. Participants completed a subset of the symptom outcome measures after completing week one and week three (weekly), and some during each ultrasound session (daily).

### Symptom outcome measures

Before any ultrasound intervention sessions, participants completed baseline surveys: Beck Depression Inventory-II (BDI-II) ^25^, PTQ ^24^, Hamilton Depression Rating Scale (HDRS) ^26^, the World Health Organization Quality of Life scale (WHO QOL)^27^, and the Columbia Suicide Severity Rating Scale (CSSRS) ^28^. These measures were re-administered following the conclusion of treatment after 1 week and 3 weeks (if applicable) of ultrasound sessions to assess weekly changes in symptoms. In addition to being administered before and after treatment, the BDI and PTQ were administered after each ultrasound session to assess daily symptom progression.

### Early remission criteria for discontinuation

To meet early remission criteria following week 1, participants must have a BDI-II score of < 13 and a HDRS score of < 8, and a PTQ score of < 18. If any of these criteria were not met, the participant continued treatment for two additional weeks.

### Remission and response criteria following treatment

After completion of the treatment protocol (i.e., after week 1 or week 3), remission (defined above) and response were assessed, with a reduction of scores to below 50% of baseline considered a response as commonly used in previous treatment literature ^15,29^.

### MRI scans

Scanning sessions included a T1 weighted structural scan, PETRA short TE scan (skull density), twelve-minute BOLD functional scan, and Susceptibility Weighted Image (SWI) before beginning ultrasound treatment, after one week of treatment, and after three weeks of treatment (if applicable). The T1 scans were used for localization and targeting and the SWI images were assessed by board-certified neurologists to assess micro-hemorrhaging. Other MRI acquisitions are not analyzed here and will be reported in a separate paper.

### Adverse events

Before each ultrasound session, subjects were asked whether they experienced adverse events that may be due to the ultrasound. For reported events, the onset and duration of the event were noted, the severity was rated, and the relationship to study procedure was assessed. After each ultrasound session, participants completed a “sensation questionnaire” to assess sensations subjects may have experienced from the ultrasound, including: itching, heat/burning, tingling, vibrating/pulsing, sound, tension, and pain. Additionally, SWI MRI images were collected at baseline and after treatment conclusion to provide an objective index of whether ultrasound may have created any damage to neurons of vasculature.

### Ultrasound session procedures

After localization and placement of the ultrasound device, each ultrasound treatment took ten minutes to complete. Participants were instructed to sit quietly, keeping their eyes open. After the ultrasound treatment was complete, the participant sat quietly for another 20 minutes, with eyes open or closed and letting their thoughts come and go.

### Ultrasound device specifications

tFUS was delivered using a custom Neuromodulation device ^30^ consisting of 128 element ultrasound array (Openwater) with the steerable ultrasound beam having the following parameters: acoustic frequency = 400 kHz, pulse duration = 5 ms, pulse repetition rate (PRR) = 10 Hz, a maximal spatial peak/temporal average acoustic intensity = 670 mW /cm^2,^, peak negative pressure 820 kPa. The ultrasound probe was secured by a custom-designed headset created by Openwater. Localite Neuronavigation Software (TMS Navigator 3.3 adapted for ultrasound device) and hardware registered the position of the probe with respect to the patient’s structural MRI, providing information to develop a novel electronically-steered, stereotactic tFUS treatment plan to the personalized target for each participant’s left anterior-medial prefrontal cortex (amPFC; MNI Coordinates −5, 45, −3^10,31,32^).

### Ultrasound targeting precision

The ultrasound array in the custom headset was affixed at the general location of the amPFC target (MNI coordinates: −5, 45, −3) with precise targeting achieved by electronic steering within limits that meet safety parameters for ultrasound exposure^30^ (Figure 1). A multi-foci, radial pattern approach was used that distributed the delivered energy in five sub-foci within 5mm from each other (which is the width of the focus in the nominal place, as defined by the −6dB pressure region). The K-Wave modeled peak energy delivery relative to the target location was highly accurate, with the −3dB centroid location of the focus falling within 1.0 +/− 1.1mm of the data measured with a hydrophone in a water tank (.02 +/− .276 mm in the lateral-axial plane, and .87 +/− 1.2mm in the axial direction). The actual pressure values estimated in K-wave and measured in the water tank agreed within 3.6 +- 1.2% within the −6dB contours.

**Figure 1.**
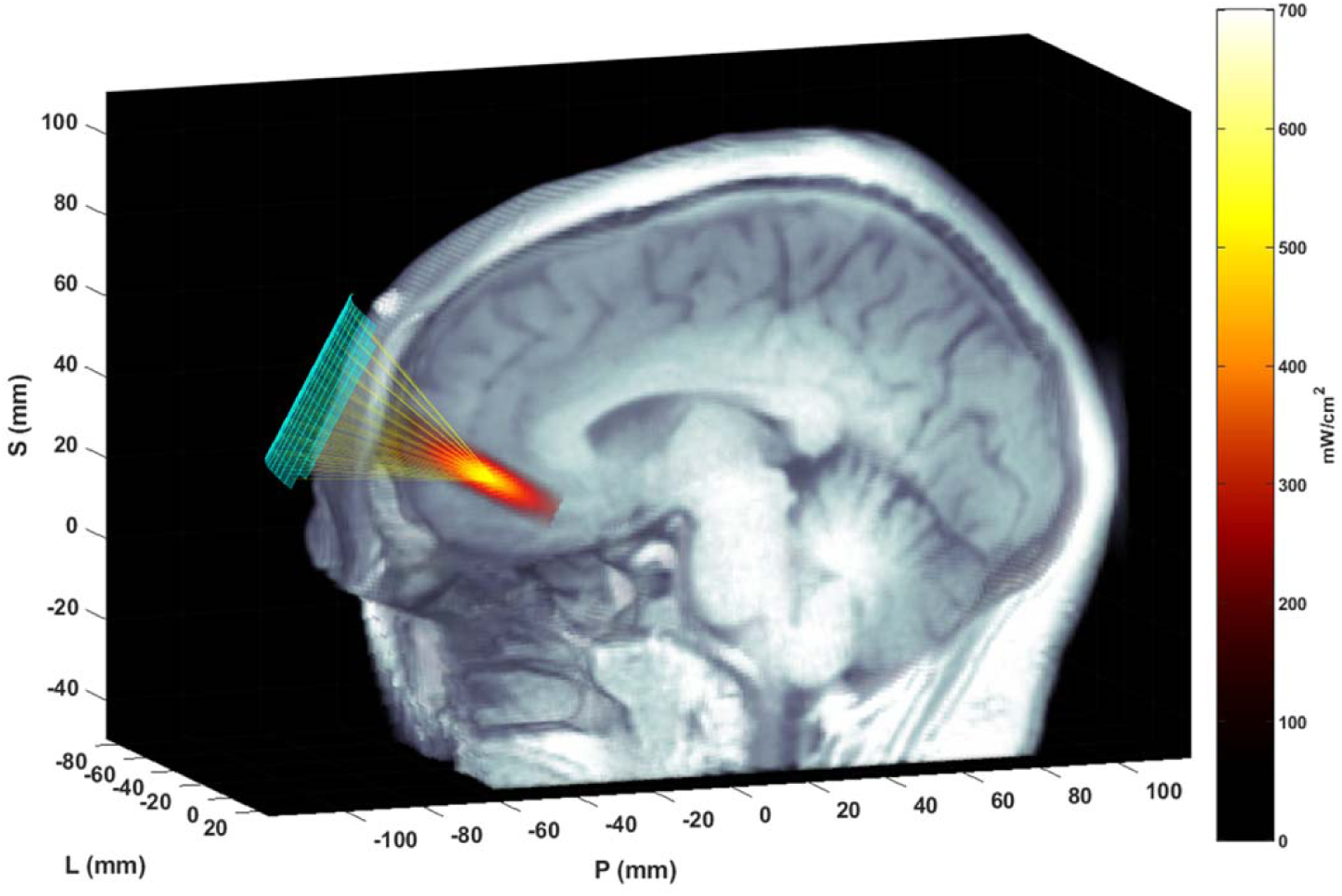
Ultrasound focusing to the amPFC. The matrix array transducer is positioned on the forehead and focuses sound through the skull and to the target. The transducer position is measured with the Localite TMSNavigator Neuronavigation system (Localite GmbH, Bonn, Germany). A focal spot, modeled based on the computed time delays using the ultrasound simulation package K-Wave, is overlain on the MRI image, representing the pulse-averaged spatial distribution of applied acoustic intensity.

### Statistical Analysis and Results

For all statistical analyses, an alpha of 0.05 was employed and significance tests were two-tailed.

### Sample characteristics

From among 386 individuals initially contacted, 247 completed the initial pre-screen web-based survey. Eighty-six potential participants completed a phone screen to confirm responses on the pre-screen survey related to eligibility, and 35 completed the Statistical and Clinical Interview for the DSM-5 to confirm a diagnosis of current depression and an absence of Mania/psychosis. Twenty participants were enrolled in the study (CONSORT diagram in Figure 2). Participant demographics are presented in Table 1. This relatively young (mean 30.4 years ± 10.0) and predominantly female (75%) sample had moderate to severe depression (BDI-II = 38.9 ± 9.3, HDRS = 19.9 ± 6.3, PTQ = 144.4 ± 6.2). The sample was also highly comorbid, and more than half had early onset depression (before the age of 13). Fifty percent of participants were currently taking medication related to their anxiety and/or depression during the intervention.

**Figure 2.**
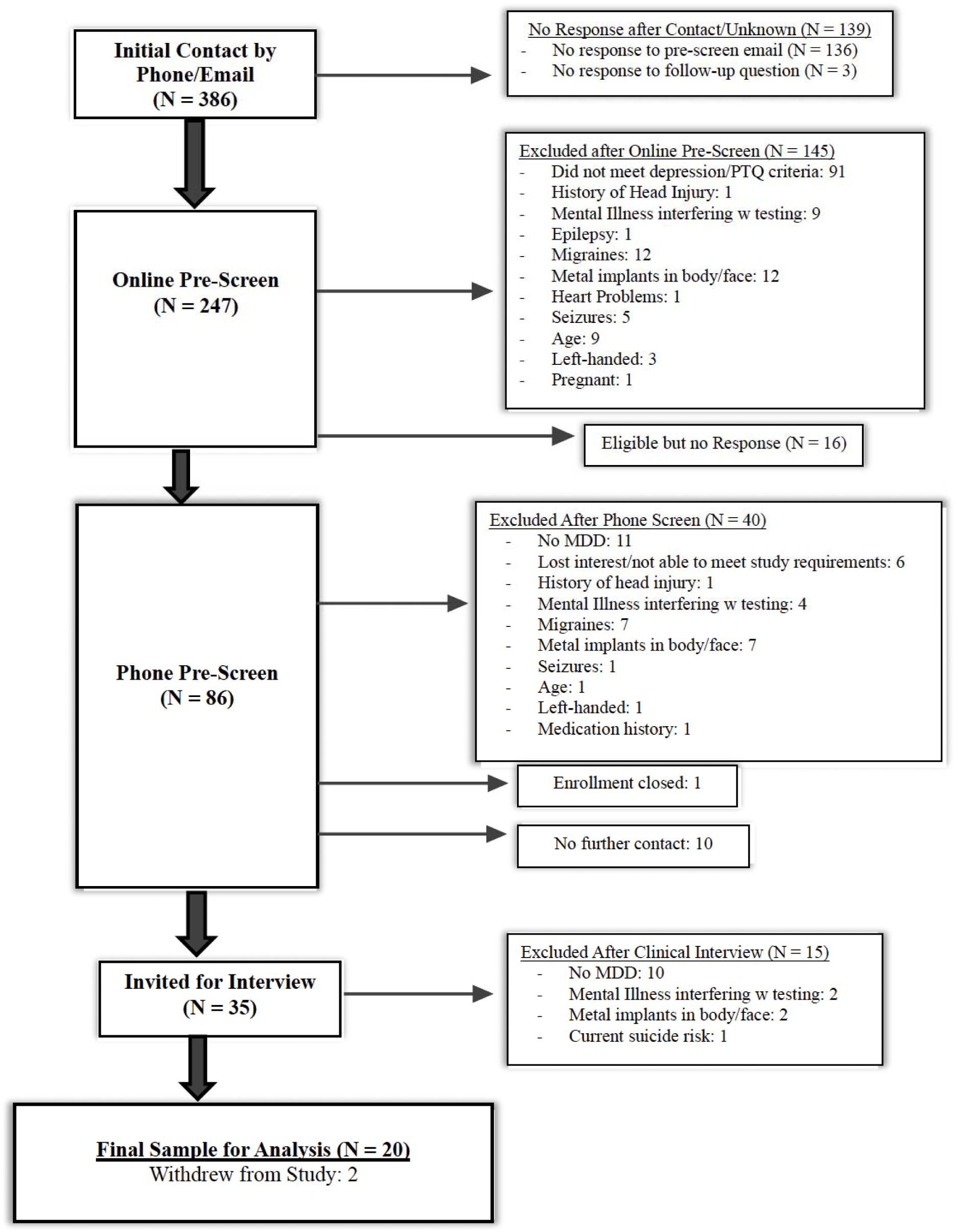
CONSORT Diagram.

**Table 1.** Participant Demographics.

| **Demographics** |  | **N = 20** |
| --- | --- | --- |
| Age, Mean (SD) |  | 30.35 (10.04) |
| Gender (F/M/Other), No. % |  | 75 / 20 / 5 |
| Years of education, Mean (SD) |  | 13.83 (1.93) |
| Race, No. % |  |  |
|  | White | 45 |
|  | Black | 10 |
|  | Chinese | 5 |
|  | Middle Eastern | 5 |
|  | Indian | 5 |
|  | Unknown | 30 |
| Ethnicity, No. % |  |  |
|  | Hispanic | 0 |
|  | Non-Hispanic | 70 |
|  | Unknown | 30 |
| Employment, No. % |  |  |
|  | Full-time | 15 |
|  | Student | 15 |
|  | Part-time | 45 |
|  | Unemployed | 25 |
| Baseline BDI-II, Mean (SD) |  | 38.85 (9.34) |
| Baseline PTQ, Mean (SD) |  | 44.35 (6.24) |
| Baseline HDRS, Mean (SD) |  | 19.90 (6.34) |
| Depression onset (Early/Teen/Adult), No. % |  | 55/ 25 / 20 |
| Comorbidities, No. % |  |  |
|  | Anxiety and Stress-related Disorder | 85 |
|  | Trauma-related Disorder | 15 |
|  | Attention Deficit Hyperactivity Disorder | 35 |
|  | Eating Disorder | 5 |
|  | Persistent Depressive Disorder | 55 |
| History of Suicidal Ideation (Passive/Active/None), No. % |  | 30 / 60/ 10 |
| Hospitalization History (Any), No. % |  | 35 |
| History of Suicide Attempts (None/One/Multiple), No. % |  | 70 / 15 / 15 |
| Current Treatment (Medication/Psychotherapy/ None), No. % |  | 50/ 20/ 10 |
| Past Treatment (Medication/Psychotherapy/ None), No. % |  | 75 / 60 / 10 |
| Current Medication Type, No. % |  |  |
|  | SSRI (Luxov, Prozac, Sertraline) | 15 |
|  | SARI (Trazadone) | 5 |
|  | NDRI (Wellbutrin) | 10 |
|  | Anti-convulsant (Gabapentin, Lamotrigine) | 15 |
|  | Beta-Blockers (Propranolol) | 5 |
|  | CNS stimulant (Adderall, Vyvanse) | 10 |
|  | Sedative (propofol) | 5 |
|  | Anti-hypertensives (Clonidine) | 10 |

### Adverse events

Dropout rate, as one index of the acceptability of tFUS treatment, was low: 10% (2/10) did not complete treatment, discontinuing after week one of treatment due to lack of symptom improvement. Dropout was not due to adverse events.

No adverse events were reported. Reported sensations (itching, heat/burning, tingling, vibrating/pulsing, sound, tension, and pain) are presented in Table 2; for aversive sensations, the modal and median endorsement was 0 (no sensation). All means were below 2.2 on the 10-point scale. For pain and tension specifically, individual reports attributed the pain and tension to the tightness of the headset, not the ultrasound itself.

**Table 2.** Sensation intensities reported on the Sensation Questionnaire.

| SENSATION | MODE | MEDIAN | MEAN | STD<br>DEV | MIN | MAX |
| --- | --- | --- | --- | --- | --- | --- |
| PAIN | 0 | 0 | 0.91 | 1.76 | 0 | 7 |
| ITCHING | 0 | 0 | 0.28 | 0.82 | 0 | 7 |
| HEAT/BURNING | 0 | 0 | 0.65 | 1.14 | 0 | 5 |
| TINGLING | 0 | 1 | 0.87 | 1.62 | 0 | 8 |
| VIBRATING/PULSING | 0 | 0 | 1.20 | 1.64 | 0 | 8 |
| SOUND | 0 | 0 | 1.36 | 1.92 | 0 | 10 |
| TENSION | 0 | 0 | 1.63 | 2.16 | 0 | 8 |

SWI images acquired at baseline before tFUS sessions and again after week 1 and week 3 were read by two board-certified neuroradiologists. SWI images are sensitive to vascular micro-hemorrhages. All 20 scans per timepoint were determined to be normal with no findings on SWI, indicating that there were no microhemorrhages resulting from tFUS delivery. Three participants’ baseline SWI readings revealed nonspecific white matter hyperintensities which may be seen with chronic microangiopathic ischemic changes and decreased susceptibility which may be related to microhemorrhages. With no change in the pre and post treatment MRI scans of these presumed microhemorrhages, they were deemed chronic.

### Outcomes

For the BDI-II and HDRS, respectively, 60% and 45% of all 20 participants met response criteria. Thirty-five percent (7/20) met remission criteria for both the BDI-II and HDRS. A multi-level model (MLM) assessed decreases in depression symptoms and RNT over the course of treatment. Each model, for each outcome of interest, BDI-II, PTQ, or HDRS score, fit “time” as the independent variable and as a random slope. A full information maximum likelihood estimator and Satterthwaite degrees of freedom adjustment were applied to each model. Given that “time” was already on a scale from 0 – 2, centering was not required. Bootstrap confidence intervals were used to demonstrate robustness to the small sample size and to account for the considerable variability in depression symptoms, while relying on a non-parametric approach that does not hinge on stringent assumptions about the underlying data distribution.

Significant decreases in depression severity and RNT were observed (Figure 3). tFUS significantly decreased BDI-II and HDRS total scores by 11.3 (p < 0.001, CI = −14.68, −8.15) and 4.3 (p < 0.001, CI = −6.21, −2.43), respectively, across time. tFUS also significantly decreased in PTQ total scores by 8.53 (p <0.001, CI = −11.01, −5.79).

**Figure 3.**
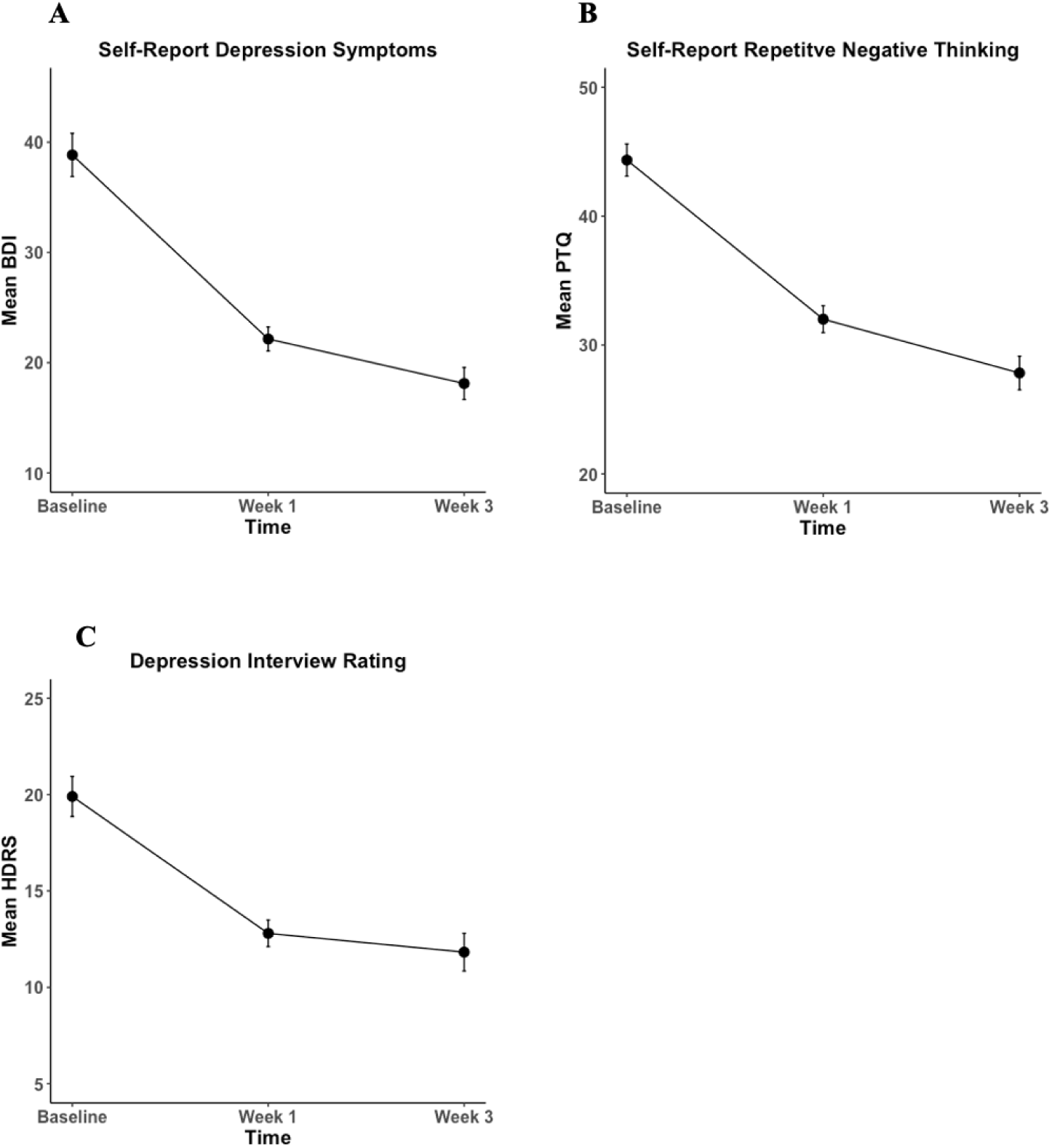
Reductions in depression symptoms and repetitive negative thought. Significant decreases in depression symptoms and repetitive negative thought over the course of transcranial focused ultrasound treatment, assessed by (A) Beck-Depression Inventory – II (BDI-II), (B) Perseverative Thinking Questionnaire (PTQ), and (C) Hamilton Depression Rating Scale (HDRS). Error bars represent within-participant standard error.

There was a significant positive relationship between change in depression and change in RNT (Figure 4), for both the BDI-II self-report (*R*^2^ = 0.67, F = 36.84 (1, 18), p < 0.001, CI = 0.76, 1.57) and HDRS interview ratings (*R*^2^ =0.37, F =10.59 (1, 18), p = 0.004, CI = 0.17, 0.79).

**Figure 4.**
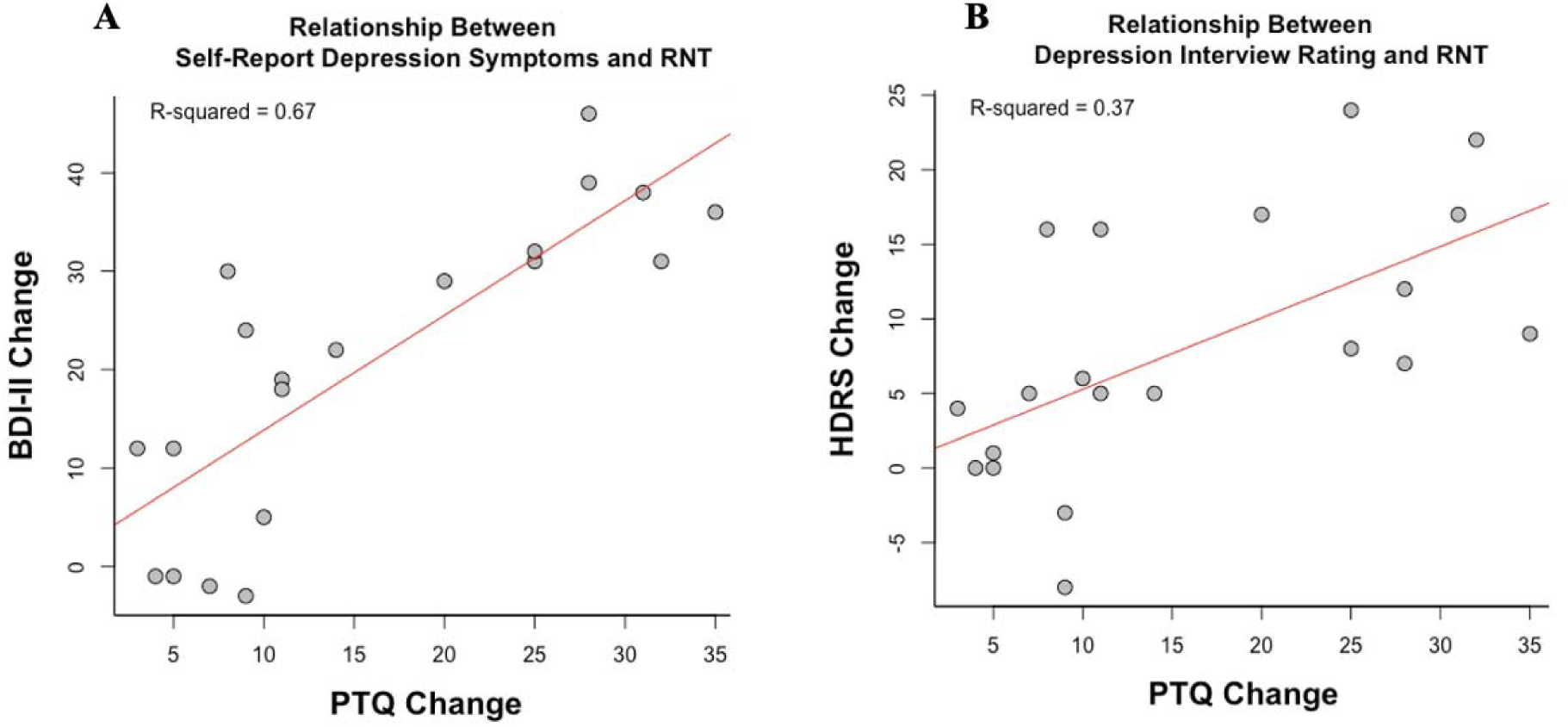
Significant associations between change in depression symptoms and change in repetitive negative thought (RNT). (A) Beck Depression Inventory – II (BDI-II) and Perseverative Thinking Questionnaire (PTQ). (B) Hamilton Depression Rating Scale (HDRS) and PTQ. The scatter plot represents a linear regression containing the R-squared value to assess the strength of the relationship and the red line to visualize the linear fit. Change scores for BDI-II, HDRS, and PTQ were computed as pre-post scores, meaning greater positive numbers reflect a greater decrease in depression severity and RNT.

### Quality of life

A multi-level model assessed changes in each of the four main WHO-QOL subscales (physical well-being, psychological well-being, social satisfaction, and environment satisfaction) over the course of treatment, with time (0 to 2) as the independent variable and random slope (Figure 5). Physical and psychological well-being significantly improved by 7.6 (p < 0.001, CI = 3.62, 11.63) and 11.9 points (p < 0.001, CI = 7.51, 16.21) and environment satisfaction increased by 5.0 (p = 0.001, CI = 2.24, 7.56). No significant improvements in social satisfaction were observed (p = 0.2, CI = −1.03, 7.44).

**Figure 5.**
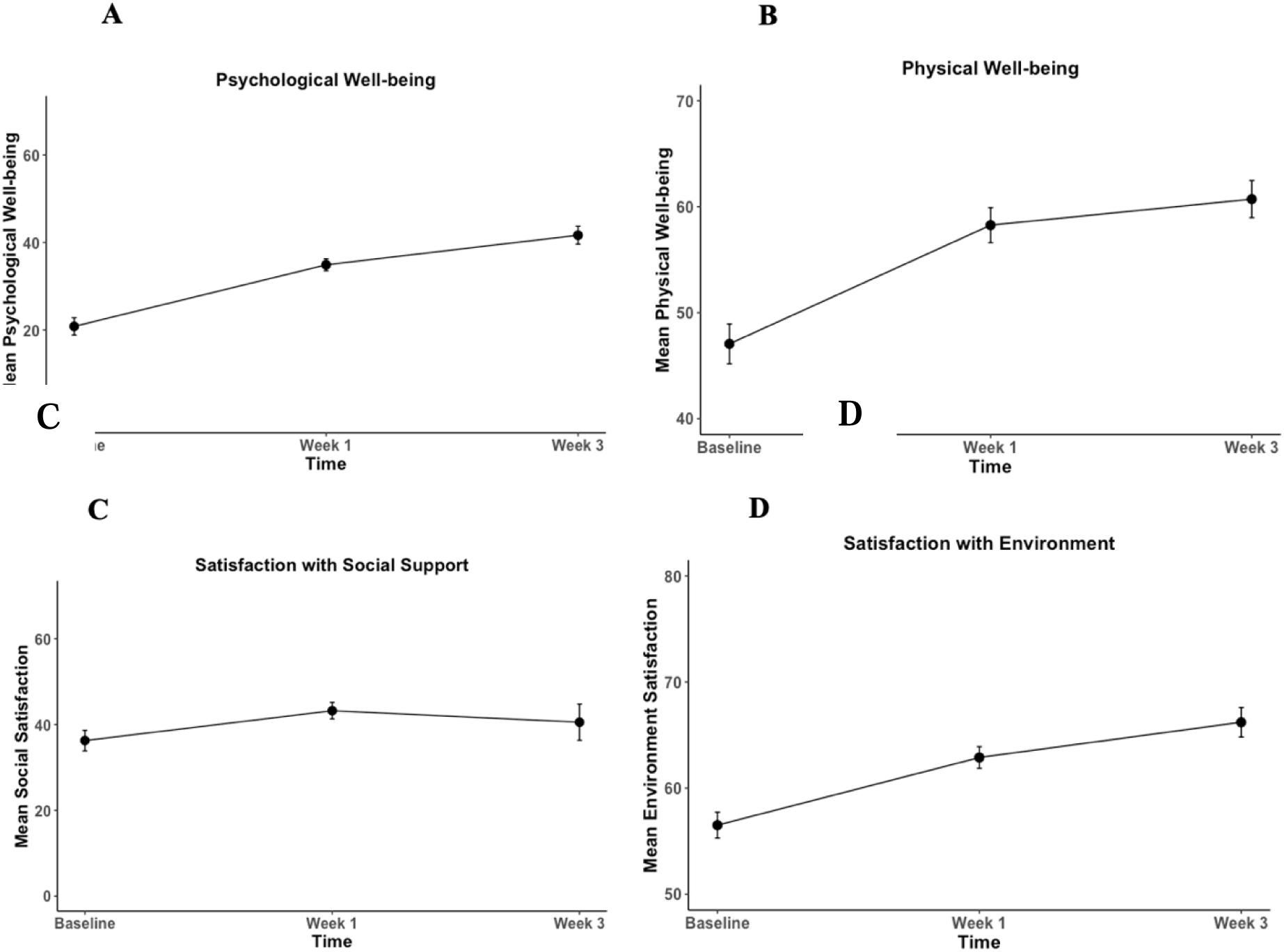
Effects of transcranial focused ultrasound on well-being using the World Health Organization Quality of Life Survey (WHO-QOL). (A) Physical Well-being. (B) Psychological well-being. (C) Social satisfaction. (D) Environment satisfaction. Significant improvement is found for panels A, B, and D. Error bars represent within-participant standard error.

## Discussion

### Adverse Events

Transcranial focused ultrasound treatment for depression using a novel, electronically-steered, stereotactic approach was successfully delivered without serious adverse events. Participants only reported transient, mild to moderate discomfort (e.g., tension and pain) which is similar to sensations experienced in many neuromodulation treatments for depression, such as rTMS ^33^. Unlike TMS or tDCS, where the source of the pain and discomfort is largely due to the delivery of the magnetic stimulation itself (e.g., skin irritation, local pain) ^23^, several participants identified the source of the pain and tension to be from the headset. Unlike other neuromodulation techniques, such as TMS, where up to 22.6% of participants experienced headaches from the active treatment ^34^, there were no reports of headaches related to tFUS delivery.

On average, previous neuromodulation techniques experience a 4.5% dropout rate due to stimulation-related adverse events ^35,36^. In the present study, zero percent of participants dropped out due to tFUS-related adverse events and only 10% of participants dropped out due to lack of positive effects of the treatment, which is also significantly better than dropout rates in traditional clinical depression trials, such as individual psychotherapy and pharmaceuticals with up to one-third drop out prior to treatment completion ^37–39^. Overall, these findings support the notion that not only is tFUS comparably safe to novel interventions such as TMS and tDCS, it may also have fewer side effects and lower dropout compared to other neuromodulation techniques.

### Reductions in depression symptoms and RNT

tFUS significantly reduced depressed mood and RNT in individuals with current major depression in just three weeks. For the BDI-II and HDRS, respectively, 60% and 45% of participants met response criteria. These percentages are comparable to traditional treatments for depression, such as antidepressants and psychotherapy (45 – 55%) in samples without substantial comorbidity; ^40^. The rates in the current study were achieved despite substantial comorbidity, a known poor prognostic sign ^41^.

An advantage of tFUS compared to traditional interventions is the rapidity of response: the response rate of 45-60% and remission rate of 35% occurred after just three weeks of treatment, which exceeds what has been found in rTMS interventions for depression with remission rates of as little as 18.6% and up to 30% after up to six weeks of treatment involving more sessions ^33,42^.The response from tFUS also occurred with fewer sessions (11) than traditional cognitive behavioral therapy^43^ (∼12 – 20 sessions, once or twice per week^44^). These findings suggest that tFUS may offer a more rapid response than traditional treatments.

### Improvements in quality of life

tFUS significantly improved physical and psychological functioning, as well as satisfaction with one’s environment. This extends previous clinical intervention work where quality of life is not commonly considered a main outcome in treatments for depression ^45,46^. Additionally, certain treatments (e.g., antidepressants) fail to lead to greater improvements in quality of life compared to controls ^47^, which prompts an important re-evaluation of what “improvement” means when developing and validating treatment protocols. It will be critical in future work to assess sustained changes in quality of life resulting from tFUS for depression, as well as treatments for depression generally.

The lack of improvement in social satisfaction after tFUS suggests the potential for future tFUS studies to augment tFUS with interventions that are known to improve social relationships and support, such as interpersonal psychotherapy and cognitive behavioral therapy ^48^, as a multimodal package that addresses the full dimensionality of improving QOL.

### Role of Repetitive Negative Thought in Depression

There was a significant, positive relationship between the change in depression symptoms and change in RNT, wherein those with greater reductions in RNT experienced greater reductions in depression symptoms. These findings support previous literature identifying the relationship between RNT and depression^8,9^, however, future work requiring larger sample sizes and a control group should aim to apply more sophisticated models coupled with longitudinal datasets to assess a predictive relationship between RNT and depression.

### Role of Default Mode Network in Depression

Our results provide preliminary support regarding the DMN’s role in depression and RNT, as we were successfully able to decrease symptoms by directly targeting a major hub of the DMN. Further evidence will include resting-state functional connectivity MRI analysis to assess whether changes in DMN connectivity track changes in depression symptoms.

### Limitations

The present study provides important evidence for the use of tFUS as a novel, targeted intervention for depression. A critical limitation is that this study was an open-label unblinded trial with a relatively small sample size. To assess whether there is a causal relationship between tFUS delivery and a decrease in depression symptoms and RNT, a randomized controlled trial with active and sham ultrasound is needed to control for nonspecific factors and minimize the impact of a placebo effect. Despite this limitation, the present findings provide a strong foundation for the implementation of tFUS as a treatment for depression with pronounced and rapid anti-depressant effects that impact quality of life, suggesting the promise of a randomized clinical trial.

## Data Availability

The data produced in the present study will not be available for the purpose of this preprint.

## Acknowledgements

This research was supported, in part, by an award from Openwater to John JB Allen. The authors are indebted to Diheng Zhang, Kelly Chen, Logan Blair, and Sarah Lass for their assistance with treatment sessions and study logistics. The authors wish to thank Jessica Andrews-Hanna for her role in target selection. Jessica Schachtner had full access to all the data in the study and takes responsibility for the integrity of the data and the accuracy of the data analysis.

